# Modeling the Post-Containment Elimination of Transmission of COVID-19

**DOI:** 10.1101/2020.06.15.20132050

**Authors:** Flávio Codeço Coelho, Luiz Max Carvalho, Raquel M Lana, Oswaldo G Cruz, Leonardo S Bastos, Claudia T Codeço, Marcelo F C Gomes, Daniel Villela

## Abstract

Roughly six months into the COVID-19 pandemic, many countries have managed to contain the spread of the virus by means of strict containment measures including quarantine, tracing and isolation of patients as well strong restrictions on population mobility. Here we propose an extended SEIR model to explore the dynamics of containment and then explore scenarios for the local extinction of the disease. We present both the deterministic and stochastic version fo the model and derive the ℛ_0_ and the probability of local extinction after relaxation (elimination of transmission) of containment, ℙ_0_. We show that local extinctions are possible without further interventions, with reasonable probability, as long as the number of active cases is driven to single digits and strict control of case importation is maintained. The maintenance of defensive behaviors, such as using masks and avoiding agglomerations are also important factors. We also explore the importance of population immunity even when above the herd immunity threshold.

## 1. Introduction

The COVID-19 pandemic is among the top three biggest in the last on hundred years, reaching levels only previously seen in Influenza pandemics[1]. At the time of writing, more than 8 million confirmed cases have been reported globally, with more than 500 thousand deaths [2]. Most affected countries still observe transmission, even if number of new cases show signs of reduction. Even with few cases, if cities (or countries) decide to lift quarantine measures, transmission may increase again due to the presence of sizeable portion of susceptible individuals suddenly at greater risk of infection.

The initial containment response varied considerably across countries, with some countries displaying more success than others in avoiding infections and subsequent deaths[3, 4, 5]. The economic impact of the containment efforts in the form of quarantines, lock-downs, suspension of international travelling and other drastic measures, has been remarkable [6]. This economic strain has forced many countries towards an early suspension of many of the most severe containment measures such as quarantines and mobility restrictions [7]. Naturally, the re-normalization of social interaction brings with it may concerns about the potential for a second wave of transmissions, which could potentially grow out of control [8].

Thus, an important question that emerges after a period of isolation is: *What’s the probability that local infections will be eliminated once the isolation is lifted?*. Evaluating scenarios for incidence evolution after these initial containment efforts requires models which accommodate both biological and population-level dynamics. In particular, models that can represent properly the immunological aspects of COVID-19 progression as well as the impact of containment mechanisms, such as quarantine and social distancing.

Many models have been proposed recently to deal with the temporal evolution of the epidemic[9, 10, 11], but one key aspect that must be considered is the contribution of stochastic fluctuations to the interruption of local transmission after the number of active cases is brought close to zero. Kucharsky et al. (2020, [12]), used an stochastic transmission model to estimate the daily reproduction number, *ℛ*_*t*_, which is often used to predict disease extinction (ℛ_*t*_ *<* 1), but empirical estimates of basic reproduction numbers are very sensitive to noise in the testing rates as well as to changes in case definition. Moreover, in real populations, *ℛ*_*t*_ can move back above one quite easily in response to changes in the population protective behavior.

In this paper, we approach the issue of local disease extinction by calculating the probability of extinction of the disease as a stochastic epidemic process. In the context of epidemics, the correct epidemiological terminology for the stochastic extinction is the local “elimination of infections”. In this paper, however, we shall continue to use the term extinction throughout, as it is shorter and more in line with the literature on stochastic processes. The probability of extinction is greater when the number of infected is low, so we calculate this probability assuming post-containment scenarios where cases have been dropped to very low numbers. We start by presenting the deterministic version of the model and derive its basic reproduction number. Then we derive an stochastic version of the same model and use it to calculate an analytical expression for the extinction probability. We conclude by looking at scenarios of local extinction and discussing how it applies to real scenarios, including also the impact of the fraction of the population already immunized upon the lifting of containment.

## 2. Methods

### 2.1. The SEIAHR model

First, we describe the (deterministic) model used to represent COVID-19 dynamics. The Susceptible-Exposed-Infectious-Removed (SEIR) model is a classic model for diseases for which it is important to take into account an incubation period, and variants of it have been employed in numerous COVID-19 modelling studies [13, 14]. Here, we propose a variation of this model with added asymptomatic, hospitalized compartments and a quarantine mechanism (fig. 1). Ther is no explicit compartments for Quarantied an dead individuals as they are pure sink states that do not influence the main dynamics.

**Figure 1:**
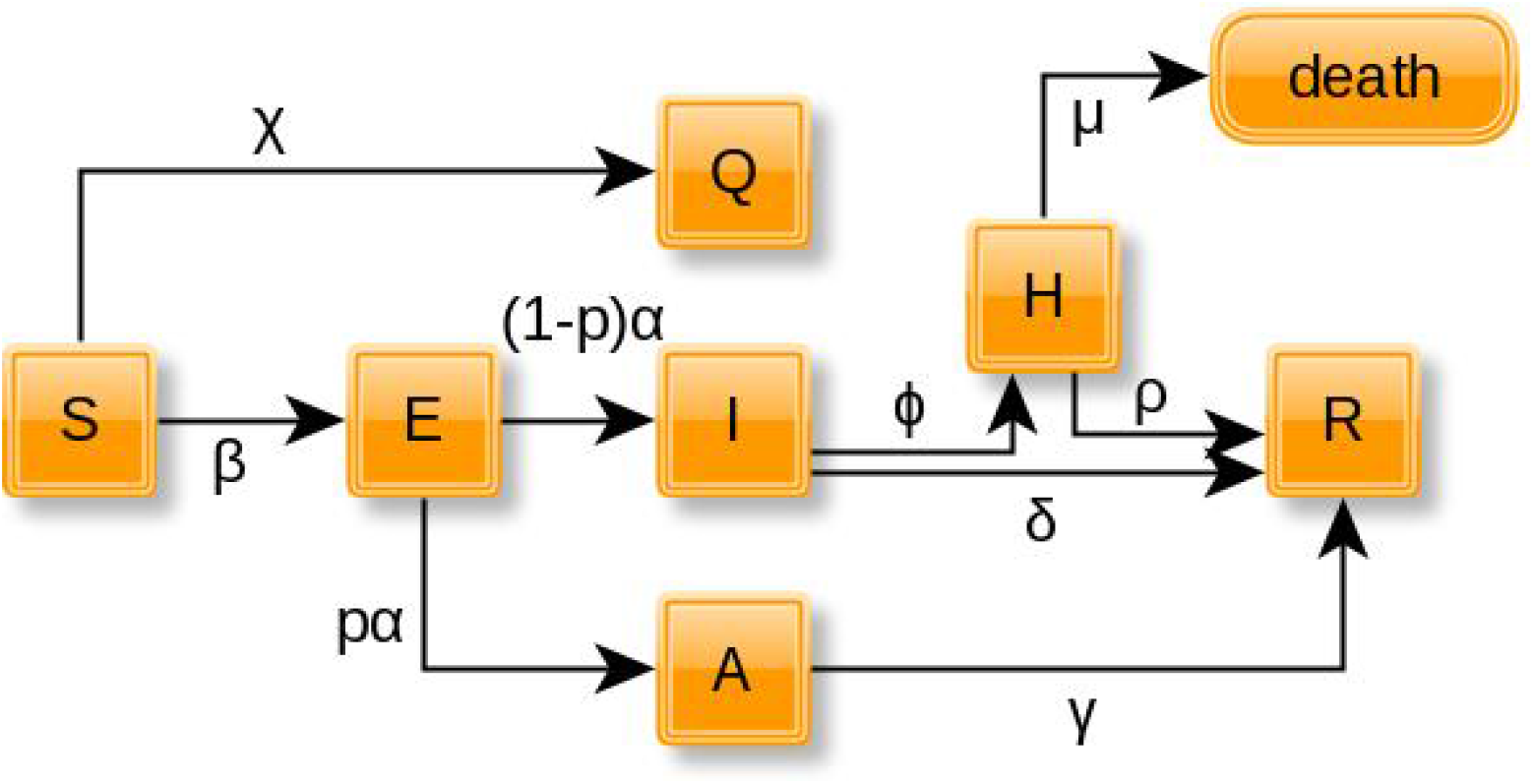
Block diagram of the SEIAHR model. compartments *Q* and *death* are included for illustrative purposes only.

The model is represented as system of ordinary differential equations:

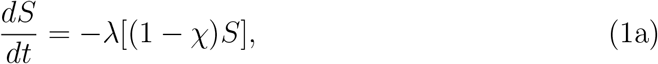

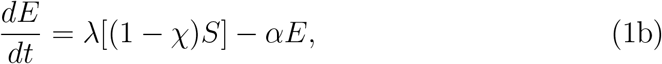

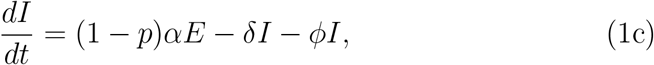

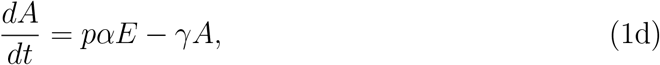

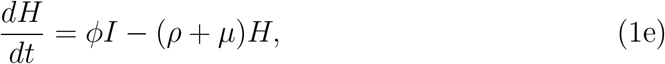

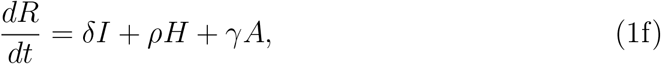

with *λ* = *β*(*I* + *A*) as the force of infection. State variables *S, E, I, A, R, H* represent the fraction of the population in each of the compartments, thus *S*(*t*) + *E*(*t*) + *I*(*t*) + *A*(*t*) + *H*(*t*) + *R*(*t*) = 1 at any time *t*. Quarantine enters the model through the parameter *χ* which can be taken as a constant or as a function of time, *χ*(*t*), that represents the modulation of the isolation policies. Quarantine works by blocking a fraction *χ* of the susceptibles from being exposed, i.e., taking part on disease transmission. Time-varying quarantine is achieved through multiplying *χ* by activation (eq. 2) and deactivation (eq. 3) functions:

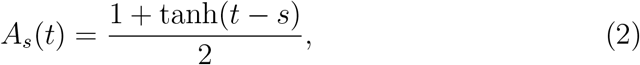

and

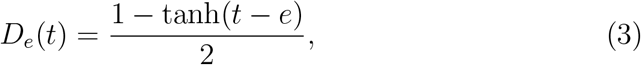

where *s* and *e* are the start and end of the isolation period (*e > s*), respectively. A finite period *τ* = *e* − *s* of quarantine can be defined by the combined effect of both functions:

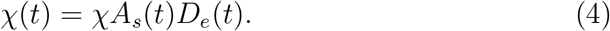

Figure 2 illustrates the activation and deactivation of quarantine.

**Figure 2:**
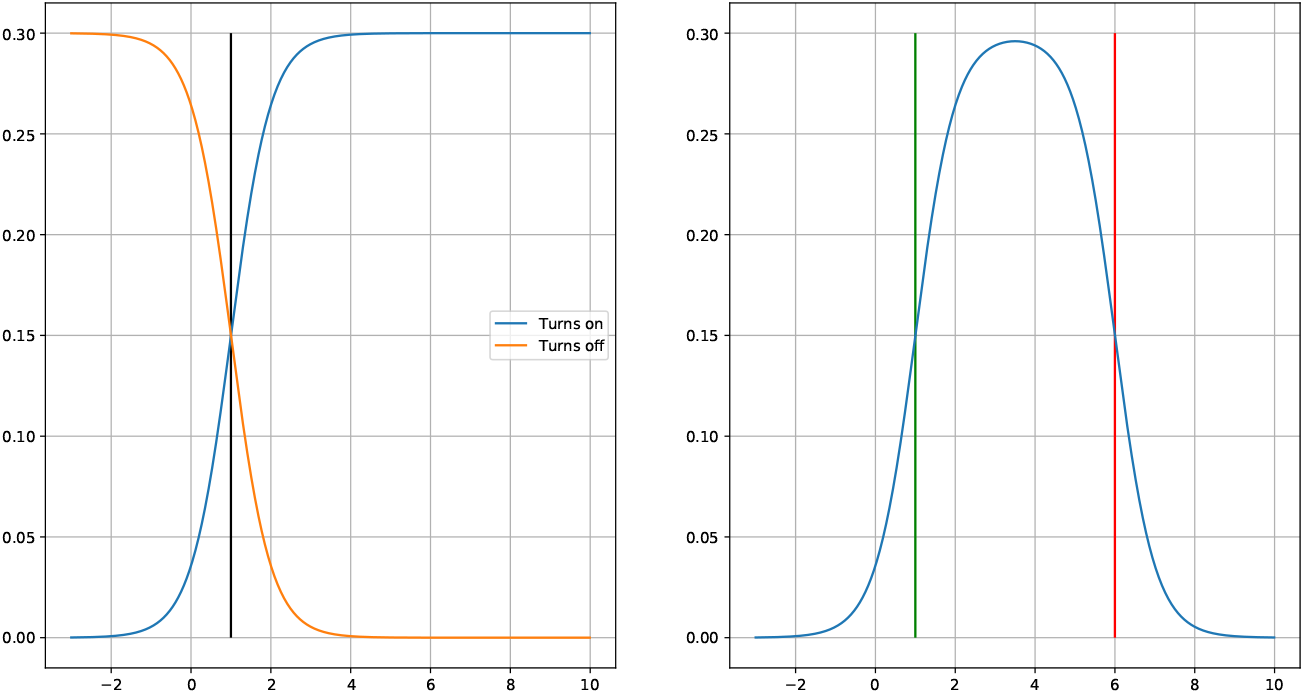
*χ*(*t*) upon activation and deactivation of quarantine for *χ* = 0.3. Panel on the left shows each function separately(*χA*_1_(*t*) and *χD*_1_(*t*)) set to *t* = 1. The right panel shows a combination with activation on *t* = 1 and deactivation on *t* = 6 (*χ*(*t*) = *χA*_1_(*t*)*D*_6_ (*t*)).

### 2.2. Basic Reproduction Number

The basic reproduction number for the SEIAHR model can be derived using the next generation matrix method[15], which we show in Appendix A. The expression for *ℛ*_0_ is

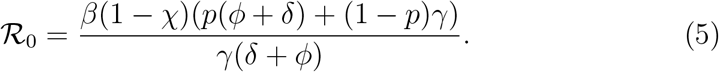

From equation (5) we can obtain *ℛ*_*t*_ = *ℛ*_0_*S*(*t*) and another reproduction number, denoted by ℛ_*c*_, which we shall call *control reproduction number*, as it represents the average number of secondary cases infected by primary cases under some control scenario – i.e. whenever *χ >* 0.

### 2.3. Probability of extinction

The question of the local extinction the disease can be more realistically addressed with a stochastic version of the SEIAHR model, where the combination of a discrete state and stochastic state transitions allow for actual extinctions to occur.

The transition rates from the ODE model (Eqs 1) listed on table 1, can be used to build a continuous–time markov chain model where time is continuous but the state is discrete, allowing for a more realistic description of population changes over time.

**Table 1:** State transitions and rates for the stochastic SEIAHR model. In the state transition column, only the changing state variables are indicated.

| Description | State transition | rate |
| --- | --- | --- |
| Infection | $(S, E) \rightarrow (S - 1, E + 1)$ | $\lambda(1 - \chi)S$ |
| Exposed to I | $(E, I) \rightarrow (E - 1, I + 1)$ | $(1 - p)\alpha E$ |
| Exposed to A | $(E, A) \rightarrow (E - 1, A + 1)$ | $p\alpha E$ |
| Hospitalization | $(I, H) \rightarrow (I - 1, H + 1)$ | $\phi I$ |
| Recovery of I | $(I, R) \rightarrow (I - 1, R + 1)$ | $\delta I$ |
| Recovery of A | $(A, R) \rightarrow (A - 1, R + 1)$ | $\gamma A$ |
| Recovery of H | $(H, R) \rightarrow (H - 1, R + 1)$ | $\rho H$ |
| Death of H | $H \rightarrow H - 1$ | $\mu H$ |

This model is a multivariate stochastic process *{S*(*t*), *E*(*t*), *I*(*t*), *A*(*t*), *H*(*t*) *}* where *R*(*t*) = *N* − (*S*(*t*) + *E*(*t*) + *I*(*t*) + *A*(*t*) + *H*(*t*)). We will leave the equation of R out, because it is decoupled from the rest of the system. A joint probability function is associated with the set of random state variables, *{S*(*t*), *E*(*t*), *I*(*t*), *A*(*t*), *H*(*t*)*}*,

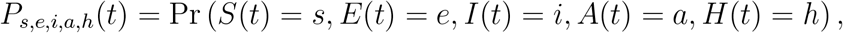

which leads to the Kolmogorov forward equation

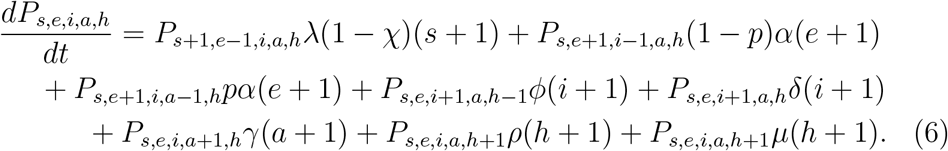

As a continuous–time branching process, the extinction threshold for the stochastic model is closely related to the corresponding one in the deterministic model but depends on the initial number of infectious individuals[16]. Based of the properties of this kind of stochastic processes, Whittle (1955) calculated the probability of extinction for the stochastic SIR model to be 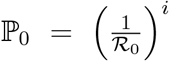, where *i* is the initial number of infectious individuals[17]. We can apply the same technique described for the stochastic SEIR model by Allen and Lahodny [16] to derive the probability of extinction for the SEIAHR model.

The analysis presented in Allen and Lahodny [16] assume proximity to the DFE with a large enough number of susceptibles and a small number of infectious individuals. Assuming a value for ℛ_0_ from other epidemics or estimated from initial exponential growth.

For a realistic application to the COVID-19 epidemic at the moment *t* of the relaxation of population lockdown, we need to acknowledge that *S*(*t*) *< N*, whilst not knowing what the exact value of *S* at time *t* and thus the effective reproduction number at the time. Nevertheless, we know from equation (5) that ℛ_*t*_ is a function of *S*(*t*). Therefore, we calculate ℙ_0_ for various values of *ℛ*_*t*_.

#### Analytical expression for ℙ_0_

The probability of extinction, ℙ_0_, can be computed analytically following the derivation of probability-generating functions for the system of equations in (6), the details of which are given in Appendix B. The probability of extinction is computed from the fixed points of the PGFs, *q*_1_, *q*_2_ and *q*_3_, which lie in (0, 1)^3^. With the fixed points in hand, we arrive at

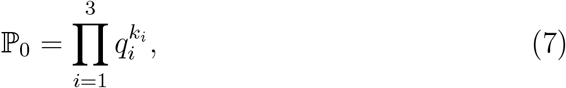

where *k*_*i*_ are the initial states *k*_1_ = *E*(0), *k*_2_ = *I*(0) and *k*_3_ = *A*(0). Here we denote the moment of relaxation of containment as *t* = 0.

#### Numerical approximation to ℙ_0_

The expression in (7), while very useful, is derived from the assumption that the initial state is small compared to the size of the population and that *S*(0) ≈ *N*. Due to the possible deviations from the theoretical value of ℙ _0_ when *S*(0) *< N*, In the results, we always present estimates of ℙ _0_, obtained by simulation as well.

We can approximate ℙ _0_ for a given ℛ_0_ and an initial number of infected individuals (*E* + *I* + *A*). This can be accomplished by running a large number of simulations of the stochastic model and computing the fraction of simulations in which the virus is driven out of the population (*E* + *I* + *A* = 0) without first causing an outbreak. To facilitate setting up the simulation to specific *ℛ*_0_, we can rewrite the force of infection by replacing *β* as a function of ℛ*R*_0_ (eq. A.8):

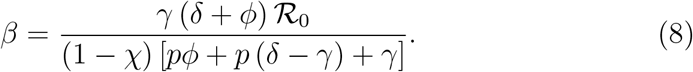

### 2.4. Effect of Increasing Seroprevalence

After a first wave of infections a fraction of the population will become immunized against SARS-COV-2. This will have a protective effect on the population even if *S*(*t*) is still above the so-called “herd immunity” threshold of 1*/ℛ* _0_. The resulting adjusted ℙ_0_ for different levels of population immunization can be determined through simulation of the stochastic SEIAHR model (eq. 6).

## 3. Results

Figure 3 shows how imposing strong containment measures can reverse the growth of the number of cases. But a second wave starts immediately after the restrictions are relaxed. Figure 4 shows that in three out of ten post-containment simulations, the containment resulted in the extinction of the disease, with the other resulting on a second wave.

**Figure 3:**
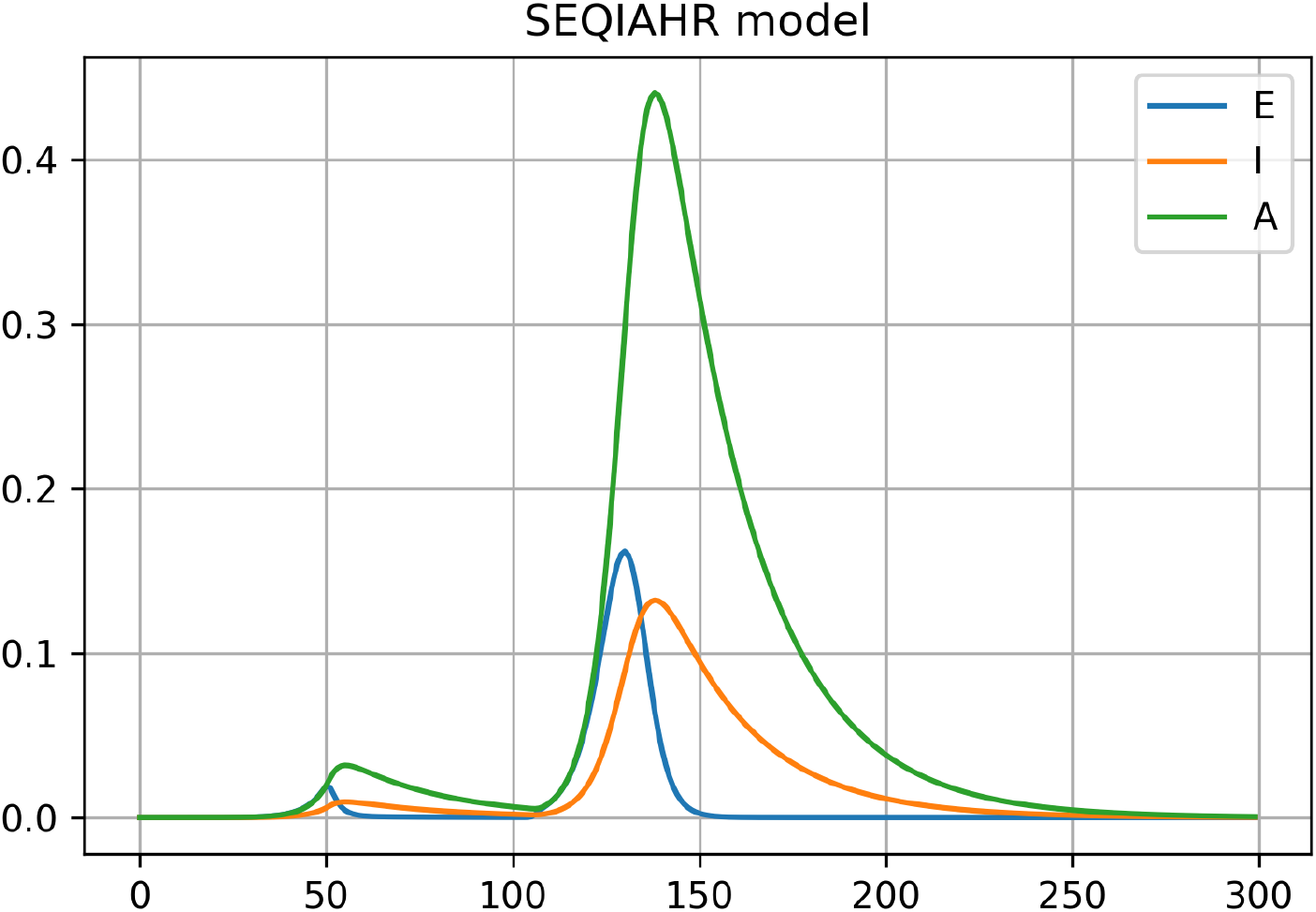
Effect of containment, *χ* = .99 for 55 days starting on the 50th day of the epidemic. Simulation parameters were ϕ = 0.01, ℛ_0_ = 1.7, *ρ* = 0.21, *δ*= 0.04, *α* = 0.34, *µ* = 0.02, *p* = 0.76, *s* = 50, *e* = 105.

**Figure 4:**
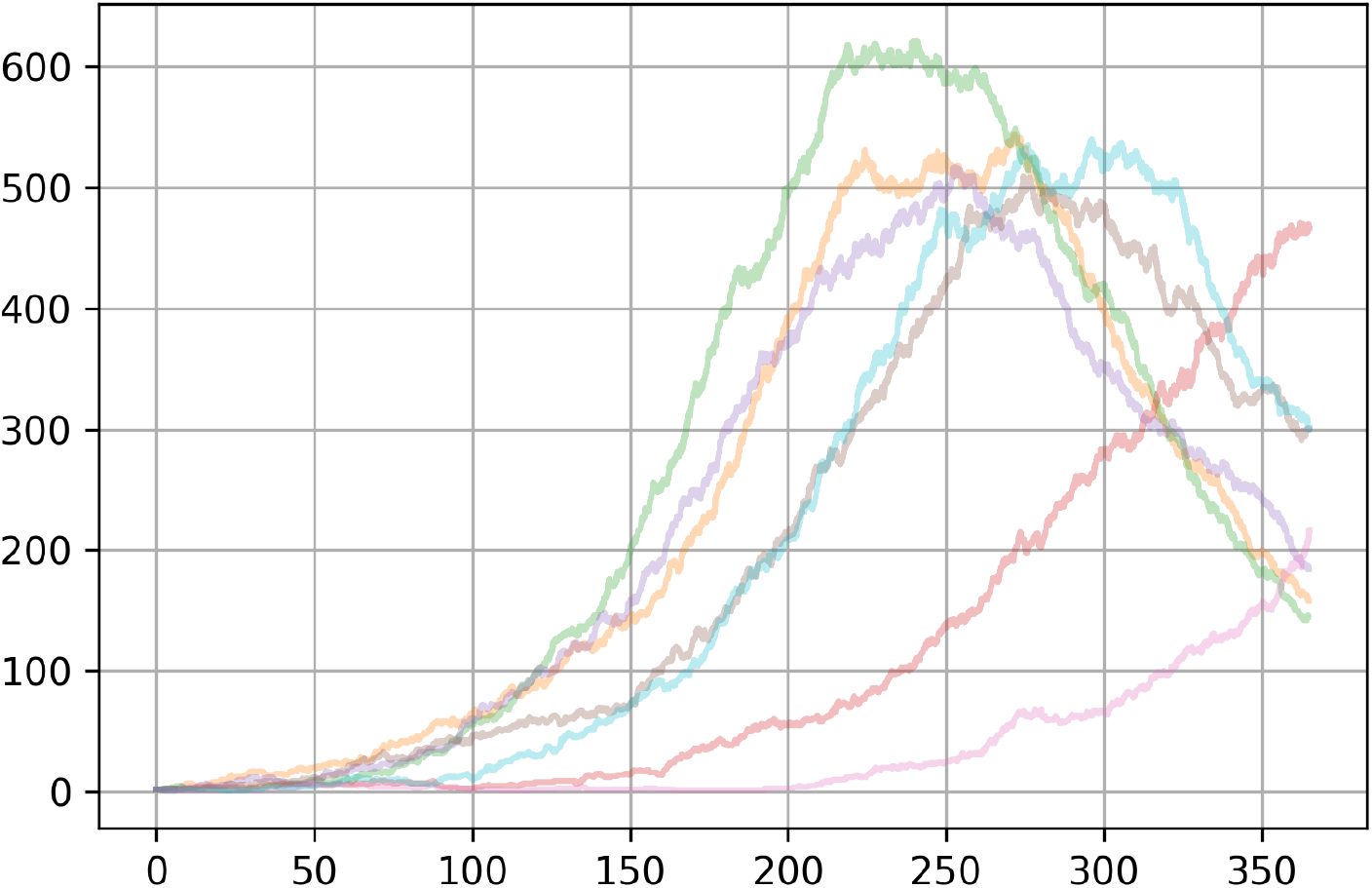
Ten runs of the stochastic SEIAHR model with the same parameters used on fig. 3. In 3 out of 10 runs the containment could eliminate the disease but the remaining 7 gave origin to a second wave. All simulations had *I*(0) = 2 and ℛ_0_ = 1.7, on a population of 5000.

Depending on the value of ℛ_*t*_ at the moment the quarantine is lifted and the number of remaining active cases, the probability of extinction can favor the stochastic extinction of the disease (Fig 5).

**Figure 5:**
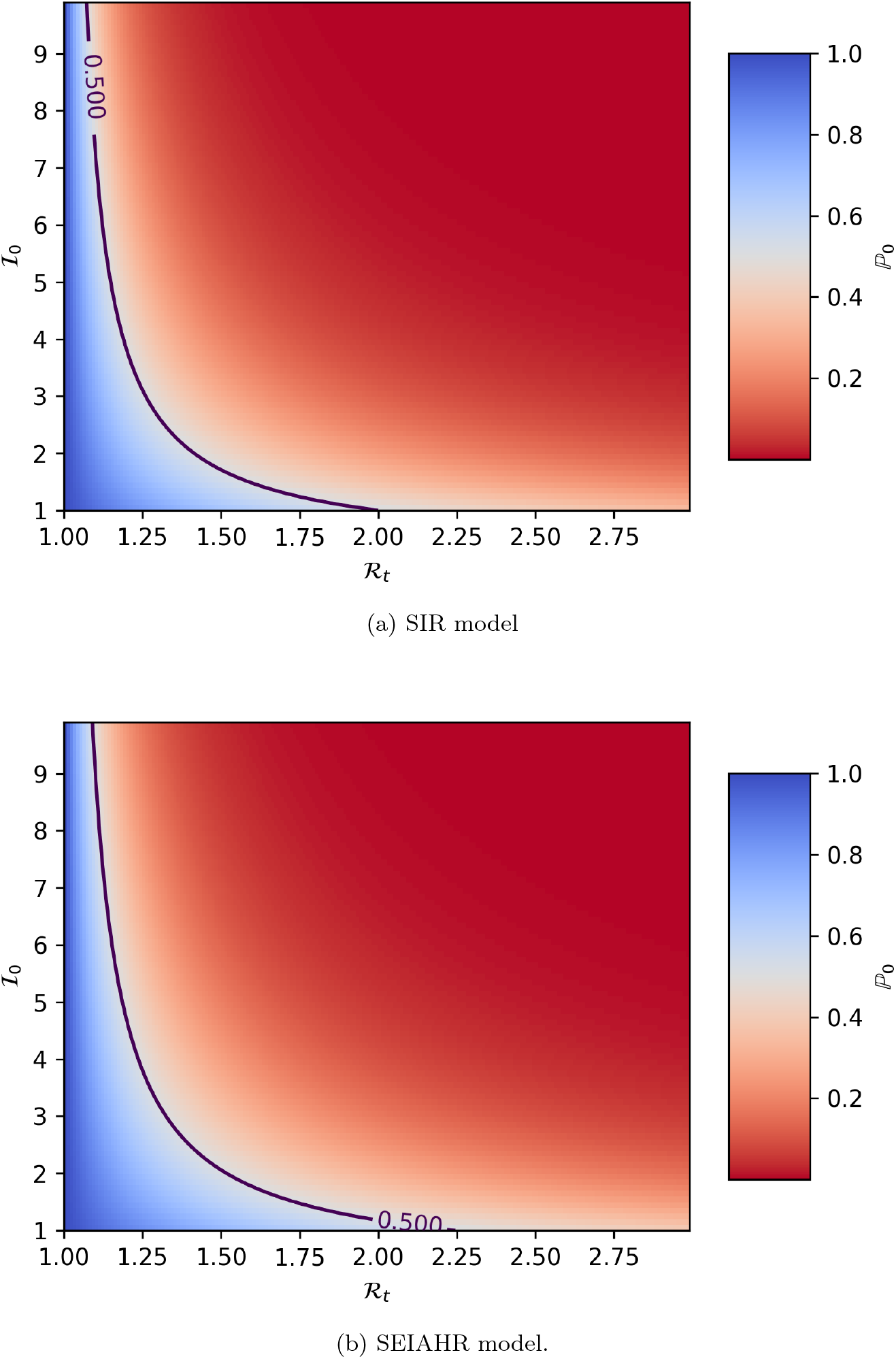
Probability of local extinction, ℙ_0_ as a function of ℛ_*t*_ and *I* _0_ .(a)for the SIR model and (b) for the SEIAHR model.

Tables 2 and 3 contain the values of ℙ_0_ for different scenarios of number of infectious, with high and low ℛ_0_ respectively.

**Table 2:** Probabilities of extinction for ℛ_*t*_ = 2.5. Approximate ℙ _0_ was calculated from a set of 10000 runs of the stochastic SEIAHR model with different number of initial infectious individuals(*I* _0_).

| $I_0$ | $E_0$ | $A_0$ | **Approx.** $\mathbb{P}_0$ | **SEIAHR** $\mathbb{P}_0$ | **SIR** $\mathbb{P}_0$ |
| --- | --- | --- | --- | --- | --- |
| 1 | 0 | 0 | 0.47 | 0.45 | 0.4 |
| 2 | 0 | 0 | 0.22 | 0.20 | 0.16 |
| 3 | 0 | 0 | 0.10 | 0.09 | 0.06 |
| 4 | 0 | 0 | 0.024 | 0.04 | 0.025 |
| 5 | 0 | 0 | 0.005 | 0.01 | 0.01 |

**Table 3:**
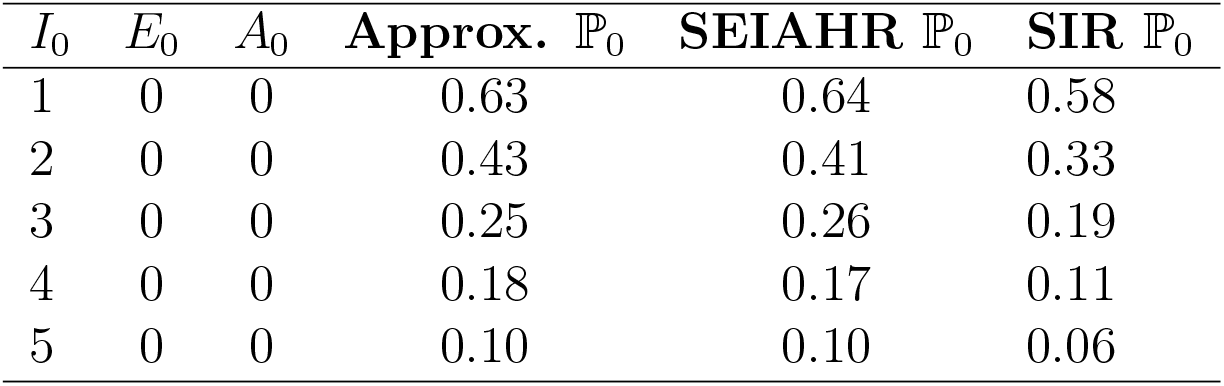
Probabilities of extinction for ℛ_*t*_ = 1.7. Approximate ℙ _0_ was calculated from a set of 10000 runs of the stochastic SEIAHR model with different number of initial infectious individuals(*I*_0_).

**Table 4:** Probabilities of extinction for ℛ_*t*_ = 1.1. Approximate ℙ _0_ was calculated from a set of 10000 runs of the stochastic SEIAHR model with different number of initial infectious individuals(*I*_0_).

| $I_0$ | $E_0$ | $A_0$ | **Approx.** $\mathbb{P}_0$ | **SEIAHR** $\mathbb{P}_0$ | **SIR** $\mathbb{P}_0$ |
| --- | --- | --- | --- | --- | --- |
| 1 | 0 | 0 | 0.91 | 0.92 | 0.90 |
| 2 | 0 | 0 | 0.82 | 0.86 | 0.82 |
| 3 | 0 | 0 | 0.75 | 0.79 | 0.75 |
| 1 | 1 | 1 | 0.71 | 0.76 | — |
| 4 | 0 | 0 | 0.69 | 0.73 | 0.68 |
| 1 | 3 | 0 | 0.66 | 0.69 | — |
| 5 | 0 | 0 | 0.63 | 0.68 | 0.62 |

We also estimated the ℙ_0_ adjusted for different fractions of immune individuals (*R*) in population from 10000 simulations of the stochastic model. The results are presented in table 5.

**Table 5:** Adjustedℙ_0_ for different levels seroconversion (*R*(0)) of the population. Results are the fraction of stochastic extinctions in 10000 simulations, with *ℛ*_0_ = 1.1.

| $I_0$ | $R(0)$ | $\mathbb{P}_0$ |
| --- | --- | --- |
| 10 | 0 | 0.35 |
| 10 | 0.1 | 0.60 |
| 10 | 0.2 | 0.82 |
| 10 | 0.3 | 0.93 |
| 15 | 0.3 | 0.91 |
| 20 | 0.3 | 0.88 |
| 45 | 0.3 | 0.78 |

Table 6 shows the expected time in days until extinction under different levels of population immunity and post-containment number of infected.

**Table 6:** Expected time to elimination of infections, in days, counting from the end of containment. Time is expressed as mean and 95% interval. Values where estimated from a 10000 runs of the stochastic SEIAHR model and rounded to the nearest integer number of days.

| Immune fraction | $I_0$ | Time to extinction in days |
| --- | --- | --- |
| 0 | 10 | 149 [38, 325] |
| 0 | 5 | 105 [17, 313] |
| 0 | 1 | 36 [1, 185] |
| 0.2 | 10 | 145 [32, 333] |
| 0.2 | 5 | 99 [15, 313] |
| 0.2 | 1 | 36 [1, 201] |
| 0.4 | 10 | 147 [34, 301] |
| 0.4 | 5 | 103 [15, 308] |
| 0.4 | 1 | 37 [1, 216] |

## 4. Discussion

Most countries who managed to control the first wave of COVID-19 did it by means of imposed restrictions on human-to-human contact. Be it through quarantine, tracing and isolation, use of masks or a combination of social distancing tactics, they managed to minimize contacts and thus transmission. Here we have introduced a transmission model (SEIAHR) in both deterministic and stochastic formulation, which includes the isolation of susceptibles as a means of reducing transmission as well as different levels of infectious individuals. From these models (deterministic and stochastic versions) it is possible to explore the consequences of social distancing as well as to calculate the probability of a second wave upon the suspension of distancing behavior. Figure 3 shows how imposing strong social distancing can interrupt the transmission, bringing the prevalence to near zero. However, if steps are not taken to permanently change the way people interact in their daily routine, at work, school, public transport, etc. The reproduction number upon lifting of the restrictive regulations, will still be substantially higher than one and will drive a powerful second wave if the population is still far from herd immunity conditions.

In the deterministic version of SEIAHR, a second wave will always happen due to the asymptotic way that the number of infectious approach zero. Treating the epidemic process as the stochastic process that it actually is, we can see that the probability of local extinction post-reactivation is quite substantial (fig 5). In the stochastic SEIAHR, extinction events will happen with slightly lower probabilities than those of a SIR model justifying using more detailed model to study this problem. Figure 4 shows stochastic extinction taking place in three out of ten runs, with ℛ_*t*_ = 1.7 and *I*_0_ = 2. The Stochastic model allows us to compute the probabilities of eliminating local transmission under various scenarios, and can be a useful tool for planning when to lift restrictions to human mobility and interaction.

Though the probabilities of elimination of local infections seem rather low on the basic scenarios described on tables 2, 3 and 4, these do not tell us the whole story. If one takes into account the immunity acquired by the population during the containment period, one can see from the results in table 5, that even if a location is still far from achieving herd immunity, any acquired immunity will greatly improve the chances local extinction substantially.

Communities that managed to contain the disease and bring it to the brink of extinction with severe economic impact, need to know how likely they are in succeeding in their fight against the disease as they return to “normal” social and economic activities. Our results also reinforce the need to run seroprevalence surveys previous to the reopening so that the probability to eliminate local infections e properly adjusted for the context of each locality. Here we explored how to improve the chances for local transmission elimination but it must be kept in mind that the ℛ_*t*_ must be kept low (through the use of masks and social distancing) in the post-containment period to guarantee elimination and even when properly executed the time to elimination can vary from weeks to months, as shown in table 6.

## Data Availability

Upon publication

## Appendix A. Derivation of the basic reproduction number

In order to use the next-generation method, we start by identifying all *m* compartments containing infected individuals. In this case they are *E, I* and *A* and *m* = 3. Let ℱ be the vector of the rates of appearance of new infections in the three infectious compartments:

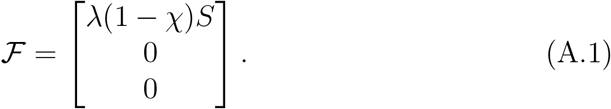

Next, we let *𝒱*_*i*_(*x*)^−^ be rate infectious individuals leave compartment *i*, and *𝒱*_*i*_(*x*)^+^ be the rate with which they enter compartment *i*:

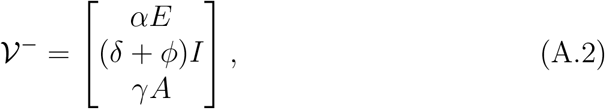

and

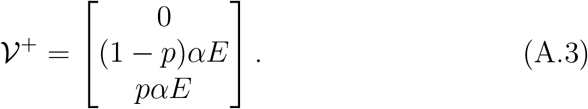

After defining these we can calculate *𝒱* (*x*) = *V𝒱*_*i*_(*x*)^−^ − *𝒱*_*i*_(*x*)^+^:

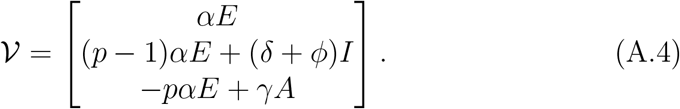

Now, if we let *x* = *{E, I, A}*and *x*0 be the Disease-free equilibrium (DFE), we can define

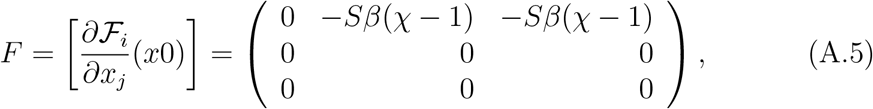

and

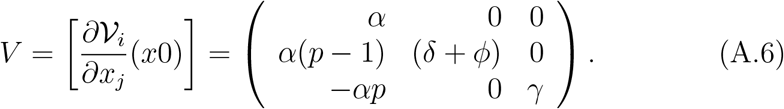

The next generation matrix is given by *FV* ^−1^:

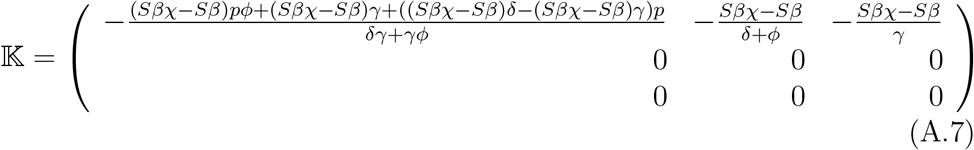

The spectral radius of 𝕂 at the DFE (when *S*(0) ≈ 1), is the basic reproduction number of the model, *ℛ*_0_ = *ρ*(*FV* ^−1^),

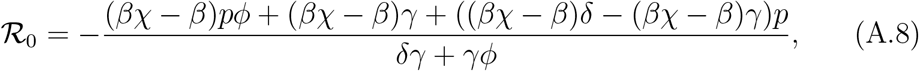

which, after simplification, gives equation (5).

## Appendix B. Probability generating functions

Following [16], we derive the probability-generating functions (PGF) for each infectious compartment. Starting with *I*_*i*_(0), the probability of an infected individual in state *i* producing offspring of type *j* given that *I*_*j*_(0) can be obtained from

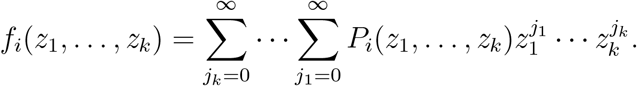

The desired probabilities can be obtained by differentiating the PGF with respect to *z*_*i*_ and setting all ***z*** to 1. Notice *f*_*i*_ has a fixed point at *z*_1_ = … = *z*_*k*_ = 1.

Computing the relevant probabilities is straightforward by keeping track of the possible transitions (given in Table 1) and considering that only one transition may occur in a given time interval Δ*t*. First, when *E*(0) = 1, *I*(0) = 0 and *A*(0) = 0, we have

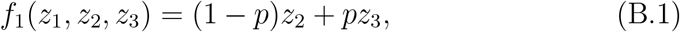

and when *E*(0) = 0, *I*(0) = 1 and *A*(0) = 0 the PGF is

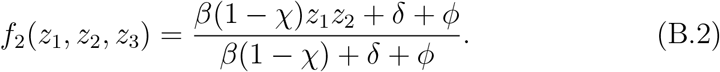

Finally, for the case *E*(0) = 0, *I*(0) = 0 and *A*(0) = 1:

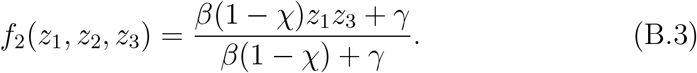

From the PGFs, we can obtain a matrix 𝕄 whose entries 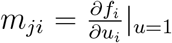 are the expected number of offspring in state *j* from an individual in state *i*. Given the PGFs above, we arrive at

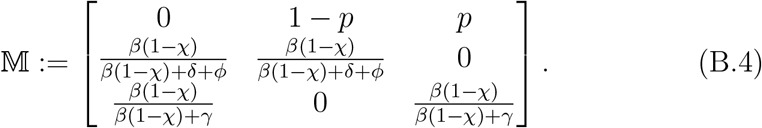

To obtain the probability of extinction, ℙ_0_, we need to find the fixed points of the PGFs, i.e. solutions to equations of the form f_i_(q_1_, q_2_, q_3_) = q_i_, q_i_ ∈ (0, 1). After some tedious algebra, we arrive at

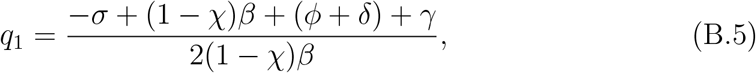

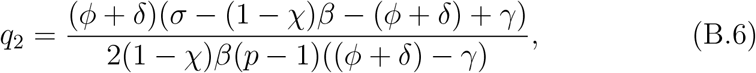

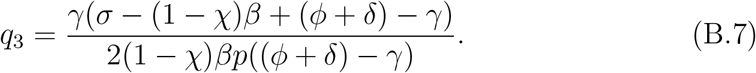

With 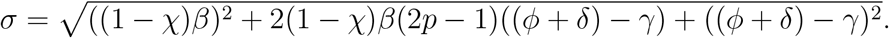.

